# What do people living with chronic pain want from a pain forecast? A research prioritisation study

**DOI:** 10.1101/2023.04.24.23289032

**Authors:** Claire L Little, Katie L Druce, William G Dixon, David M Schultz, Thomas House, John McBeth

## Abstract

**Background:** People with chronic pain report feelings of uncertainty and unpredictability around their future pain. A pain-forecasting model could provide important information to support individuals to manage their daily pain and improve their quality of life. To be useful, the model should be developed with people living with chronic pain. We conducted Patient and Public Involvement (PPI) work, with the aim of this PPI to design the content of a pain-forecasting model by (1) learning participants’ priorities in the features of pain provided by a pain forecast and (2) understanding the benefits that participants perceive they would gain from such a forecast.

**Methods:** A focus group of 12 participants identified potential features, benefits and drawbacks of a pain forecast. In a survey, participants with chronic pain (*n* = 148) prioritised the identified pain features and perceived benefits.

**Results:** Focus group participants identified anticipatory anxiety and fears around data-sharing as potential drawbacks. Survey respondents prioritised forecasting of pain flares (68%) and fluctuations in pain severity (64%). Specific priorities about pain flares were the timing of the onset and the severity. Of those surveyed, 75% would use a future pain forecast and 80% perceived making plans (e.g. shopping, social) as a benefit.

**Conclusions:** For people with chronic pain, the timing of the onset of pain flares, the severity of pain flares and fluctuations in pain severity were prioritised as being key features of a pain forecast, and making plans was prioritised as being a key benefit.

**Plain English Summary:** Chronic pain is a symptom of many long-term health conditions. People with chronic pain have reported that the severity of their pain is both uncertain and unpredictable. To combat this, we want to build a pain forecast, to predict future pain severity. We hypothesise that a pain forecast would reduce pain-related uncertainty and improve quality of life. It is important that a pain forecast provides useful information to people living with chronic pain. Therefore, this work aimed to understand why participants might use a forecast, and what they would want to see in a pain forecast.

A focus group was conducted to identify features, benefits and drawbacks of a pain forecast. A survey was then conducted to prioritise the features and benefits. Participants of the focus group highlighted concerns around data-sharing and potential anxiety about knowing when pain might happen. Survey participants prioritised a forecast that provided information about pain flares (periods of increased pain severity) and fluctuations in pain severity. The key perceived benefit of a forecast was the ability to make plans (such as shopping and social plans).

## Background

Chronic pain (i.e. pain lasting at least three months) is experienced by an estimated 43% of adults in the United Kingdom (1,2). Chronic pain conditions are associated with significant individual and societal burden. They are among the leading causes of disability globally (3). Individuals report that pain interferes with their professional and social lives, affects their relationships, and decreases their quality of life, mood and sleep (4). In the UK, 13.4% of sickness days were due to musculoskeletal conditions in 2021 (5). Although up-to-date figures are scarce, the economic costs of chronic pain are considerable. For example, back pain (a common cause of chronic pain) cost up to £12.3 billion in the UK in 1998 (6) and chronic pain conditions cost 1.5–3% of European GDP in 2012 (7).

The severity of chronic pain is a key driver of outcome, with more severe pain associated with worse outcomes including poorer physical and mental health–related quality of life (8–10), mood (11–13), and social and work participation (14,15). However, the absolute level of pain severity is not the only important driver of outcome. Variability in pain severity is also an important factor. The severity of chronic pain is not stable over time, and individuals experience intra- and inter-daily fluctuations in pain severity and pain flares which are characterised by a rapid increase in pain severity (16–22). People living with chronic pain report that the variability in pain severity is unpredictable, and this unpredictability leads to feelings of uncertainty (23,24) that permeates every sphere of their lives through a decreased ability to work, missed social events, and avoidance in making commitments (25,26). There is a clear desire to reduce the unpredictability of pain severity, with patients often asking how their pain might manifest in the future.

Pain is a complex biopsychosocial phenomenon and predicting variability in pain severity, including pain flares, will be challenging. It will involve identifying and understanding the complex relationship between time-varying biological, psychological and social exposures, discerning how those are associated with changes in pain severity over time, and developing models to forecast those changes. We propose that a personalised pain-forecasting model could reduce pain-related uncertainty by providing predictions of future pain. We have identified factors that are associated with variability in pain severity and could be used as predictors in such a model, including prior pain experience, physical activity (27,28), mood (29,30), sleep quality (31) and environmental exposures (here, the weather) (32).

Recent developments in digital data collection tools offer a solution to capturing these data. Patient-generated health data in chronic pain are already used to track daily symptoms including pain symptoms over time (33), to inform models of care (34) and to facilitate conversations between clinicians and patients (35). Other spheres have shown the feasibility of using patient generated health data to forecast symptoms. For example, individualised prediction models exist for forecasting the diagnosis and prognosis of Covid-19 (36), the presence of anxiety and depression (37), the severity of hayfever symptoms (38) and the level of physical fatigue (39). It is feasible that patient-generated health data could also be used to forecast the variability in pain severity. However, the features that a pain forecasting model should predict are not yet clear.

There are many potential pain features that could be predicted by a pain forecasting model including, for example, the level of forecasted pain severity described as an absolute value, the level of change in forecasted pain severity described as an absolute or proportional increase, the timing of that change, and the variability in pain severity over time. It is not clear which, if any, of these features people living with chronic pain would prioritise in a pain forecast. Patient and Public Involvement (PPI) is defined as work done *with* members of the public and can be conducted to involve stakeholders in the research process, including in identifying research priorities (40,41). Conducting PPI in the process of developing a pain forecast would ensure that the forecast is suited to the needs and priorities of its users (42). Thus, identifying and prioritising pain features in PPI activities forms the first stage in producing a pain-forecasting model.

The objectives of this PPI work were to design the content of a pain forecast by (1) learning participants’ priorities in the features of pain severity provided by a pain forecast and (2) understanding the benefits that participants perceive they would gain from such a forecast.

## Methods

### Overall study design

Two PPI activities were conducted with individuals with chronic pain. The first PPI activity was a focus group to inform the second PPI activity, a survey of people living with chronic pain. The aim of the focus group was to identify potential pain features that could be produced by a pain forecast and a list of potential benefits of a pain forecast. The aim of the survey was to prioritise these features and benefits in a larger sample of people living with chronic pain. These PPI activities were approved by the Proportionate University Research Ethics Committee at the University of Manchester (Ref: 2021-11862-19751). The activities are reported in line with the GRIPP2 (Guidance for Reporting Involvement of Patients and the Public) checklist (41), which is provided in Additional file 1.

### Focus group

A semi-structured focus group was conducted with individuals with chronic pain to produce a list of meaningful pain features that could be provided by a forecast and to understand the perceived potential benefits of a forecast. A focus group was chosen as it allowed us to explore reasons behind the choices and to allow participants to build on each other’s ideas (43). A single focus group was conducted due to time and budget constraints.

We sought to recruit up to 12 individuals who were at least 18 years old, who self-reported having a non-cancer chronic-pain condition, lived in the UK, and could read English. Participants were recruited through social media and newsletters of charity organisations related to non-cancer chronic-pain conditions (see Additional file 2) and shared through professional social media accounts of colleagues. Potential participants completed a screening questionnaire, providing demographic information on their gender, ethnic group, age bracket (18–25, 26–45, 46–65 or 66+), self-reported chronic-pain condition(s) from a multiple-choice list and length of time since diagnosis. Participants for the focus group were then selected using purposive sampling, ensuring variation in age, gender, ethnic group, number and type of chronic-pain condition(s) and time since diagnosis. Recruited individuals provided informed written consent and were reimbursed for their time and expenses, in line with PPI guidelines from the National Institute for Health and Care Research (44).

The focus group took place in August 2021 and lasted approximately 90 minutes. Due to the ongoing COVID-19 pandemic, the focus group was held online using Zoom. Three researchers (authors CL, KD and JM) co-facilitated the focus group and made written, anonymised field notes.

The structure of the focus group is provided in Additional file 3. Discussion topics focussed on the pain features of a forecast that participants identified as potentially beneficial, how a forecast could be used in day-to-day life and how the survey (the second PPI activity) should be structured. These discussions led to revisions of the survey, details of which are provided in the results section. The structure of the focus group included a general group-level introduction (facilitated by CL), breakout-room discussions (facilitated by CL, KD and JM), and a final group-level discussion.

Group-level discussions in the focus group were audio-recorded through Zoom, without video recordings, and subsequently transcribed verbatim by CL. Field notes from breakout rooms were made by all facilitators. Participants’ views on potential features of a pain forecast, and how a forecast may be used in day-to-day life were noted and subsequently used to inform multiple-choice questions in the survey.

### Survey

The second PPI activity was a survey of people living with chronic pain. This survey aimed to learn participants’ priorities regarding the potential features and perceived benefits of a pain forecast, using the features and benefits identified by the focus group participants.

The survey was distributed in October and November 2021. The recruitment strategy was identical to that of the focus group. Members of the focus group were not prohibited from completing the survey. Consent was provided electronically at the start of the survey, and only complete surveys were analysed. We aimed to receive at least 100 completed surveys.

The survey collected demographic information and included priority-setting questions and multiple-choice questions. The demographic information collected were participants’ gender, age, chronic pain condition(s) and site(s) of pain. Priority-setting questions asked participants to order multiple-choice options in preference order, using a drag-and-drop feature. Multiple-choice questions asked participants to select one or multiple options, with free-text space for further elaboration if required. Priority-setting and multiple-choice questions were written following discussions in the focus group, and so details are deferred to the Results section. The full survey can be viewed in Additional file 4.

Analysis of the survey questions was conducted descriptively. For priority-setting questions, the distribution of respondents prioritising each option as most important to least important was calculated. For multiple-choice questions, the number and percentage of participants selecting each response was calculated. Sensitivity analyses using chi-squared tests were conducted to compare the responses of participants with commonly reported pain conditions to those without.

Due to the small number of free-text responses, they were not directly analysed but are reported following the relevant questions.

## Results

### Focus group

Demographic data of the 12 participants are provided in Table 1. There were nine females and three males, with all age brackets (18–25, 26–45, 46–65 or 66+) represented. Six participants had been living with chronic pain for at least five years, and five participants had two or more chronic-pain conditions. Chronic-pain conditions of the participants included osteoarthritis, chronic headache, fibromyalgia, neuropathic pain, rheumatoid arthritis and spondyloarthritis.

**Table 1:** Demographics of focus group participants (*n* = 12)

| **Table 1: Demographics of focus group participants (*n* = 12)** |  |
| --- | --- |
|  | Number of participants |
| Age |  |
| 18—25 | 1 |
| 26—45 | 3 |
| 46—65 | 6 |
| 66+ | 2 |
| Gender |  |
| Female | 9 |
| Male | 3 |
| Ethnicity |  |
| Asian British | 1 |
| Black British | 1 |
| British Chinese | 1 |
| South Asian | 1 |
| White British | 7 |
| White British & Black African | 1 |
| Number of chronic pain conditions |  |
| 1 | 7 |
| 2 or more | 5 |
| Chronic pain condition* |  |
| Chronic headache | 2 |
| Fibromyalgia | 2 |
| Neuropathic pain | 2 |
| Osteoarthritis | 5 |
| Rheumatoid Arthritis | 2 |
| Spondyloarthritis | 2 |
| Other | 3 |
| Time since diagnosis |  |
| Under 5 years | 6 |
| 5 or more years | 6 |
| *Column exceeds 100% since participants can have multiple chronic pain conditions |  |

Discussions in the focus group were centred on three overarching themes: the pain features, if any, that participants wanted a forecast to provide, the perceived benefits of a forecast and the potential drawbacks of using a forecast in daily life. These themes are discussed in more detail below, followed by the implications they had on the survey questions.

The first overarching theme that participants discussed was the pain features of a forecast. Participants described pain features of interest in relation to periods of pain flares (when pain severity increased for a number of days) and periods of low or no pain severity (when pain severity decreased for a number of days). First, they spoke about the timing of these periods (such as when a period of low pain might begin), as this would have implications on activities that could be undertaken, and may impact how work and social activities could be managed in advance:

P6 (F66+): “I gave an example of a [colleague] at the moment who’s got a frozen shoulder and she’ll be coming back on a phased return but the timing of that may be helpful through the [forecast]”

Second, they discussed the duration of pain severity at high or low levels. Knowledge about the duration of pain severity at certain levels would also allow participants to plan activities and relevant interventions. For example, a participant in a breakout room commented that they may visit a chiropractor if their pain was predicted to last for a substantial period of time.

Third, participants wanted to know information about their pain-related quality of life and other symptoms (such as fatigue). Participants in breakout rooms noted that these other symptoms were not always correlated to pain severity and wanted to know information about them separately.

P9 (F46–65): “I think you should maybe focus on quality of life. We all have different levels of pain, different levels of fatigue, we are all different. But what is important for me is totally different to the next person… It’s on what you would accept as a quality of life”

The second overarching theme that participants explored were the perceived benefits of a pain forecast. The most commonly reported benefit of a forecast was that it could improve the ability to make plans, including meeting family, planning grocery shopping, planning pharmacological interventions, organising non-pharmacological interventions and adapting work:

P3 (F26–45): “For me… the biggest advantage would be planning medication… I tend to go for the lowest meds, and then regret it because I’m still in pain and, oh God, now I can’t take this, or I could take it but then I have stomach issues, all the rest of it. So… if I could get my drugs more accurate to how it’s going to be, my pain medication, my PMR [steroid medication], that would really help, I think.”

Another reported benefit was that participants hoped a forecast might support them in understanding triggers of pain, including how variables such as weather, stress and exercise might affect pain severity. In breakout rooms, participants discussed the empowerment granted through understanding their own triggers of pain.

In the third overarching theme, participants identified potential drawbacks of a pain forecast. First, participants highlighted potential mental-health challenges, such as anxiety and stress induced by having information about pain events, including pain flares:

P3 (F26–45): “There are mental health disadvantages like anticipatory anxiety if the [forecast] tells you you’re going to feel rubbish in a week.”

Among these concerns, participants voiced fears of a self-fulfilling prophecy if they expected pain severity to increase.

Other participants highlighted mental-health challenges related to inputting pain-severity scores, which may encourage higher focus on the pain that they are trying to manage.

Second, participants were anxious about the potential implications of data collection during a pain forecast and fears of data sharing with employers and government officials:

P7 (F46–65): “Who has access to the data? I think it would put a lot of people off if people thought that employers are going to have access to this data.”

P6 (F66+): Would it be “used by occupational health departments in organisations?”

Based on the discussion of the focus group, priority-setting and multiple-choice questions for the survey were written. Of the three overarching themes, questions were developed regarding the potential features and benefits of a pain forecast. As the drawbacks related to the implementation of a pain forecast, these were not included in the present survey. All suggested pain features of the focus group discussion were included, asking survey participants to prioritise the importance of timing, duration and symptoms during periods of low pain and pain flares. All reported benefits were included, asking survey participants to select which benefits applied to them regarding planning, applying pharmacological and non-pharmacological interventions and understanding triggers of pain.

### Survey

There were 148 respondents to the survey. Demographic information and data regarding chronic pain condition can be found in Table 2. Approximately 90% of respondents were female and over three quarters were aged between 36 and 65. The most commonly reported chronic-pain conditions were fibromyalgia (46%) and osteoarthritis (33%). Sensitivity analysis discovered no differences between the responses of participants with these common pain conditions and other pain conditions (see Additional file 5). Nine participants reported only ‘other’ pain conditions. These participants reported ankylosing spondylitis, psoriatic arthritis, scleroderma, systemic lupus erythematosus and juvenile arthritis.

**Table 2:** Demographics and chronic pain condition of survey respondents (*n* = 148)

|  |  |  |  |
| --- | --- | --- | --- |
|  |  | Number of participants | Percentage of participants |
| Age | 18–25 | 11 | 7.4% |
|  | 26–35 | 19 | 12.8% |
|  | 36–45 | 34 | 23.0% |
|  | 46–55 | 27 | 18.2% |
|  | 56–65 | 40 | 27.0% |
|  | 66+ | 17 | 11.5% |
| Gender | Male | 13 | 8.8% |
|  | Female | 134 | 90.5% |
|  | Non-binary/third gender | 1 | 0.7% |
| Chronic pain condition* | Fibromyalgia | 68 | 46.0% |
|  | Osteoarthritis | 49 | 33.1% |
|  | Chronic headache | 32 | 21.6% |
|  | Neuropathic pain | 28 | 18.9% |
|  | Rheumatoid Arthritis | 19 | 12.8% |
|  | Spondyloarthritis | 17 | 11.5% |
|  | Unspecific Arthritis | 12 | 8.1% |
|  | Other | 69 | 46.6% |
| *Column exceeds 100% since participants can have multiple chronic pain conditions |  |  |  |

Results of the multiple-choice question *“Which of the following would you like a pain forecast to provide for you?”* are shown in Table 3. The most commonly selected features were pain flares (68%) and fluctuations in pain severity (64%). Features of pain severity on an ordinal scale (47%) and periods of low pain (35%) were less commonly selected.

**Table 3:** “Which of the following would you like a pain forecast to provide for you?”

|  |  |  |
| --- | --- | --- |
| **Pre-specified response** | **Number of participants who selected response** | **Percentage of participants who selected response** |
| Information about a pain flare | 100 | 67.6% |
| Information about fluctuations in pain severity | 94 | 63.5% |
| Information about pain severity on a scale of 1 to 5 | 70 | 47.3% |
| Information about a period of low/no pain severity | 51 | 34.5% |
| Other/None | 13 | 8.8% |
| *Responses to the question: "Which of the following would you like a pain forecast to provide for you?" Participants could select more than one option.* |  |  |

Only two relevant free-text responses were provided to this question, both referring to fluctuations in pain severity:

“Will my future pain graphs differ from those in the past?”

“Compare it to half hour ago, a couple of hours ago, yesterday etc with a higher lower method”

Results of the priority-setting question “*If we could predict a pain flare, what specific information would you want to know?*” are shown in Figure 1. The respondents ranked six statements. Onset of a pain flare was the first priority for 92 (62%) of respondents. Severity of a pain flare was the first or second priority for 50% of respondents. Pain-related quality of life and variation in other symptoms were given fifth or sixth priority by 70% and 58% of respondents, respectively. Responses to this question highlight that the onset of a pain flare and severity of a pain flare are clear priorities for respondents.

**Figure 1:**
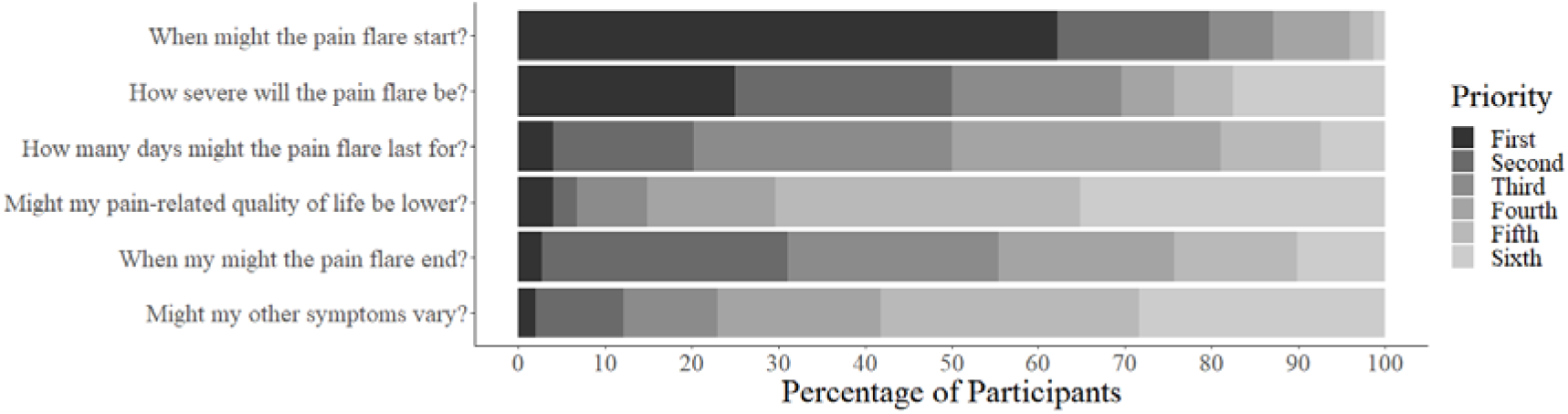
Survey respondents’ priorities of the pre-specified responses relating to pain flares. Respondents were prompted with the question: “If we could predict a pain flare, what specific information would you want to know?”. Percentages of participants ranking each statement as their first, second, third, fourth, fifth or sixth priority are reported.

In free-text responses, nine respondents highlighted that they also wanted information about the triggers of their pain flare. Specific triggers that were cited were hormonal cycles, weather, environment and mood. One participant wanted information about the acceleration of the pain flare and one wanted information about medication to take during a flare.

Results of the priority-setting question “*If we could predict a period of low pain severity, what specific information would you want to know?*” are shown in Figure 2. The respondents ranked five statements. Onset of a period of low pain severity was first priority for 70 (47%) respondents. Respondents did not show great variability among the other responses. First or second priority was given to the duration of the period by 36% of participants, to variation in other symptoms by 35% of participants, to the end of the period by 34% of participants, and to pain-related quality of life by 32% of participants. There was therefore only a clear priority for information about the onset of the period of low pain severity.

**Figure 2:**
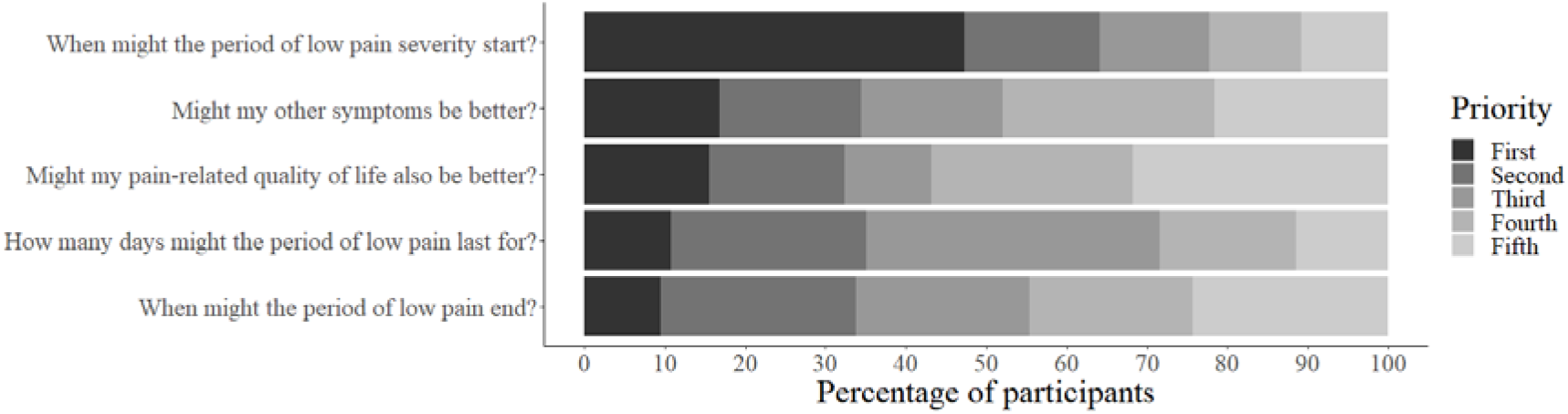
Survey respondents’ priorities of the pre-specified responses relating to periods of low pain severity. Respondents were prompted with the question: “ If we could predict a period of low pain severity, what specific information would you want to know?”. Percentages of participants ranking each statement as their first, second, third, fourth or fifth priority are reported.

Of the free text responses, eight respondents referred to wanting information about the triggers of their low pain (e.g. barometric pressure) or variables that they could control (e.g. exercise). One participant wanted to understand how typical their experience is among others, one wanted information about treatments, and one wanted information about specific days.

Participants were asked the multiple-choice question: “*If a pain forecast could provide useful information for you, do you think that you would use a pain forecast?”* (Table 4). Of those surveyed, 75% would use a pain forecast.

**Table 4:**
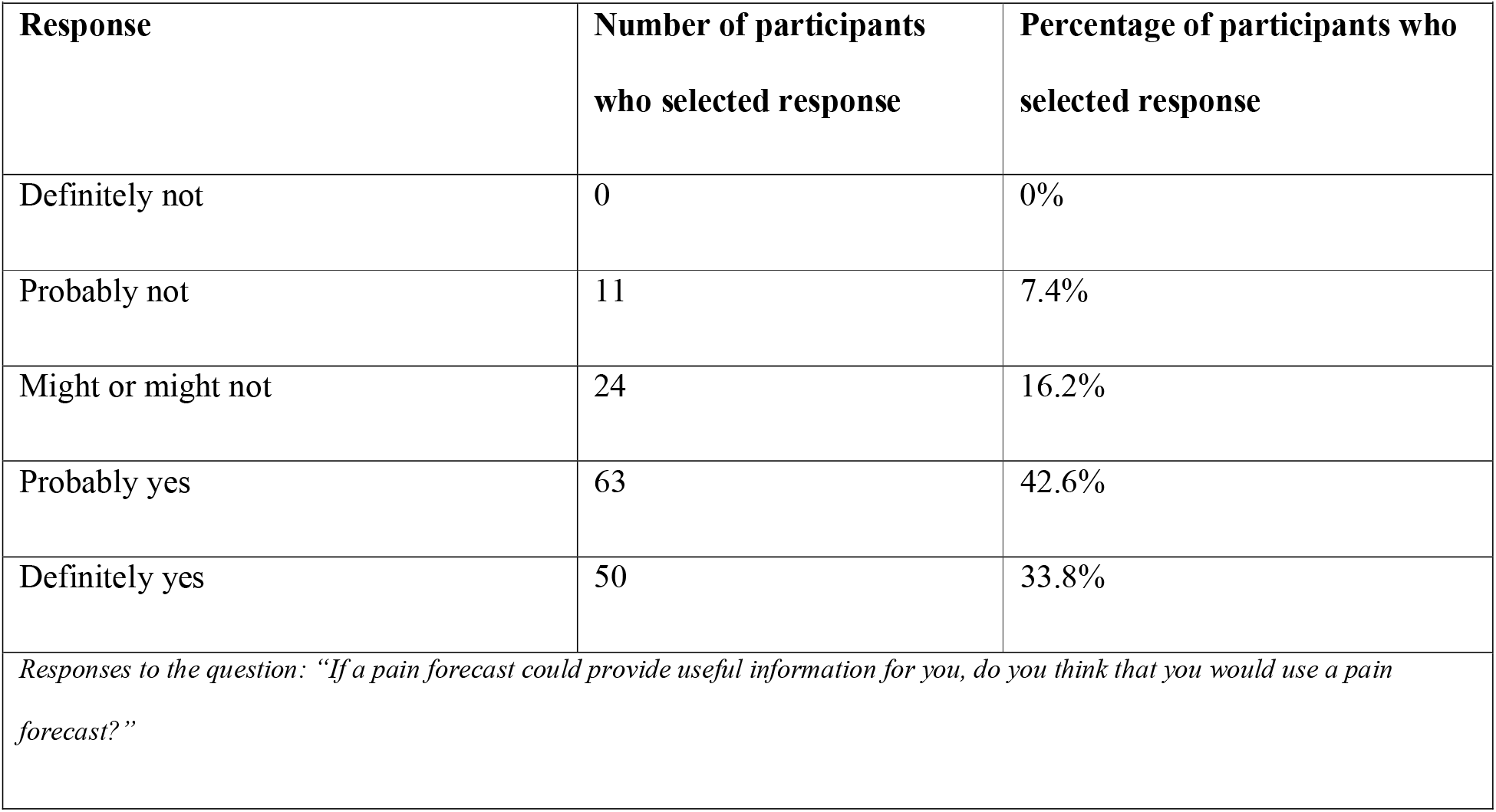
“Do you think that you would use a pain forecast?”

| **Table 4: “Do you think that you would use a pain forecast?”** |  |  |
| --- | --- | --- |
| **Response** | **Number of participants who selected response** | **Percentage of participants who selected response** |
| Definitely not | 0 | 0% |
| Probably not | 11 | 7.4% |
| Might or might not | 24 | 16.2% |
| Probably yes | 63 | 42.6% |
| Definitely yes | 50 | 33.8% |
| *Responses to the question: “If a pain forecast could provide useful information for you, do you think that you would use a pain forecast?”* |  |  |

All participants were also asked: “*What would you use a pain forecast for?”* (Table 5). The most common reasons were making plans (83%) and understanding individual triggers of chronic pain (76%). In addition, 47% and 31% of respondents would use a pain forecast to plan pharmacological and non-pharmacological interventions respectively. Therefore, making plans and understanding triggers are highlighted as the most likely benefits, although planning pharmacological and non-pharmacological interventions may also be of interest to a number of users.

**Table 5:** “What would you use a pain forecast for?”

| **Table 5: "What would you use a pain forecast for?"** |  |  |
| --- | --- | --- |
| **Pre-specified response** | **Number of participants**<br>**who selected response** | **Percentage of participants**<br>**who selected response** |
| To help make plans (e.g. shopping, social) | 123 | 83.1% |
| To understand the triggers of my pain | 113 | 76.4% |
| To know when my pain severity might be<br>better/worse | 92 | 62.2% |
| To help plan non-pharmacological<br>interventions | 70 | 47.3% |
| To help choose which medication to take | 46 | 31.1% |
| Other/None | 16 | 10.8% |
| *Responses to the question "What would you use a pain forecast for?" Participants could select more than one option.* |  |  |

Relevant free-text responses were:

“To improve my overall self management of my conditions”

“To let work know times where I might need time off to recover so that it’s not out of the blue for them and they can prepare for me to be off if I need to”

“To analyse the development of my condition”

“Exercise planning”

“To help me understand my condition more”

“To look forward to some good times!”

## Discussion

In our PPI activities, the content of a pain forecasting model was discussed with people with chronic pain in a focus group and subsequently prioritised in a UK-based survey. We report that survey participants highlighted an interest in a forecast involving periods of pain flares and fluctuations in pain severity. Within a priority-setting question about pain flares, the most highly ranked features were the timing of the start of their flare and the severity of their flare. Taken together, the participants in these PPI activities were clear of their interest in receiving information about the timing of the start of their flare, the severity of their flare, and fluctuations in their pain severity.

Over three quarters of the surveyed population reported that a forecast would help them to make plans and assist in understanding triggers of their pain. Respondents would also use a pain forecast to understand when their pain may be better or worse, to guide the use of non-pharmacological and pharmacological interventions. However, the focus group also highlighted concerns about the use of a forecast in day-to-day life, including anticipatory anxiety and sharing of personal data.

Developing the survey following the focus group ensured that the themes presented were identified as being important and meaningful to individuals with chronic pain. The survey results can also be interpreted in the context of topics raised in the focus group, including how a forecast might be used to support going back to work or to plan pharmacological interventions. As a result, the PPI activities allow a pain forecast to be developed with the feedback from stakeholders.

There are limitations to the PPI activities that should be considered. The representation of different conditions in our survey may have been impacted by the charities that shared the advertisements with their members, perhaps explaining the high prevalence of fibromyalgia among our respondents. This would impact the results if participants with certain conditions had different priorities to other people with chronic pain, and those conditions were over-represented in the surveyed population. However, sensitivity analyses among the subset of participants with fibromyalgia compared to the subset of participants without fibromyalgia, and the subset of participants with osteoarthritis compared to the subset of participants without osteoarthritis found no differences in the reported responses.

Recruitment advertisements clarified that participants would be commenting on a pain forecast, and respondents therefore had an interest in commenting on this topic. A large proportion of our survey participants were interested in a pain forecast and so the results may be less generalisable to those people who would initially be unsure or less inclined to use a forecast.

Both PPI activities recruited participants online, primarily due to the ongoing COVID-19 pandemic. A pain forecast may be implemented in a future digital intervention and users would then be required to access the internet. However, our work did not include participants who may be less inclined to use the internet for reasons including access and digital literacy. Our findings are therefore not generalisable to these populations and if a digital intervention is developed, work with these populations should be considered.

Previous work has reported on the longer-term prognosis of chronic pain. For example, some studies have followed people with chronic pain over several months or years and identified different trajectories of pain severity among people with chronic pain (45,46). Other studies have identified prognostic factors associated with chronic pain outcomes (47). However, the participants in our focus group and survey have highlighted the importance of forecasting pain on a shorter-term, to support daily activities.

The benefits of a pain forecast extend previous work. Flurey et al. (23) reported that patients expressed frustration at the unpredictability of pain flares and this led to participants cancelling or altering plans. Fullen et al. (26) also found that individuals with chronic low-back pain reported missing out on social events and avoided making commitments, due to the unpredictability of their pain. Our work highlighted that respondents would value a forecast that reduced the unpredictability of their pain, particularly around the timing and severity of pain flares. Our participants highlighted the importance of making plans as a key benefit of a forecast, likely due to the frustration previously reported which has resulted in avoidance of making plans.

The drawbacks highlighted are also consistent with previous work. Among the challenges in collection and analysis of patient generated health data, privacy concerns have previously been highlighted (48), in line with concerns of focus group participants. Any future mobile application of a pain forecast should follow standards of privacy and security (49) and clearly communicate these to users. Furthermore, as pain is widely accepted within the biopsychosocial model (50), and rumination and catastrophizing are associated with increased pain severity (51), concerns around anticipatory anxiety should be thoughtfully considered.

## Conclusions

These PPI activities indicated a high level of interest in a pain forecast and our participants were clear that pain features should include the timing of the start of a pain flare, the severity of a pain flare, and fluctuations in pain severity. Future work will develop a statistical pain forecasting model, to predict these identified features. As one of the key benefits of a pain forecast is the identification of triggers, a future model should be interpretable by its users. Drawbacks highlighted in the focus group, such as the impact of anticipatory anxiety should also be considered during the production of a forecast. Wider interest will be determined in the future, based on the uptake of a forecast and continued involvement of stakeholders and evaluation of a forecast will ensure that priorities indicated by participants translate into real value.

## Data Availability

The datasets generated and analysed during the current study are not publicly available as data were collected for patient and public involvement activities and consent was obtained for the sharing of anonymous quotes and aggregated data only. However, data are available from the corresponding author on reasonable request.

## List of Abbreviations

PPI: Patient and Public Involvement

## Declarations

### Ethics approval and consent to participate

This study was approved by the Proportionate University Research Ethics Committee at the University of Manchester (Ref: 2021-11862-19751). Informed written consent was provided by members of the focus group. Informed electronic consent was provided by respondents to the survey. This procedure to take informed consent electronically from the respondents was approved by the Proportionate University Research Ethics Committee at the University of Manchester. All methods were carried out in accordance with relevant guidelines and regulations (Declaration of Helsinki).

### Competing interests

WGD has received consultancy fees from Google, and DMS has received consultancy fees from Palta, both unrelated to this work. All other authors have no conflicts of interest to declare.

### Funding

This work was supported by infrastructure support from the Centre for Epidemiology Versus Arthritis (grant number 21755).

TH receives funding from the Royal Society (grant number INF/R2/180067) and the Alan Turing Institute for Data Science and Artificial Intelligence.

### Authors’ contributions

All authors contributed to the conception of the study. CL, KLD and JMcB designed the study and conducted the focus group and survey. CL analysed the results and wrote the first draft of the article. All authors contributed to revising the final article.

## Acknowledgements

We are grateful for the contributions of the members of the Patient and Public Involvement focus group and the respondents to the survey.

## Notes

### Author Declarations

The Proportionate University Research Ethics Committee at the University of Manchester (Ref: 2021-11862-19751) gave ethical approval for this work.

