## Additional File 1 for "What do people living with chronic pain want from a pain forecast? A research prioritisation study"

Additional file 1: GRIPP2 short form

| **Section and topic** | **Item** | **Reported on page No** |
| --- | --- | --- |
| 1: Aim | Report the aim of PPI in the study | 5 |
| 2: Methods | Provide a clear description of the methods used for PPI in the study | 6–8 |
| 3: Study results | Outcomes—Report the results of PPI in the study, including both positive and negative outcomes | 9–14 |
| 4: Discussion and conclusions | Outcomes—Comment on the extent to which PPI influenced the study overall. Describe positive and negative effects | 14 |
| 5: Reflections/critical perspective | Comment critically on the study, reflecting on the things that went well and those that did not, so others can learn from this experience | 14–15 |

GRIPP2 short form from: Staniszewska S, Brett J, Simera I, Seers K, Mockford C, Goodlad S, et al. GRIPP2 reporting checklists: tools to improve reporting of patient and public involvement in research. BMJ. 2017 Aug 2;j3453.
