## Additional File 2 for "What do people living with chronic pain want from a pain forecast? A research prioritisation study"

Additional File 2: Charity organisations that advertised the study

| Charity organisation | Method of advertisement |
| --- | --- |
| Action for ME | Website, Twitter, Facebook |
| Arthritis Action | Website |
| Arthritis and Musculoskeletal Alliance (ARMA) | Newsletter, Twitter |
| Backcare | Website |
| Lupus UK | Online forum |
| Migraine Trust | Website, Twitter, Facebook |
| National Axial Spondyloarthritis Society (NASS) | Website, Newsletter |
| National Rheumatoid Arthritis Society (NRAS) | Website, Twitter, Facebook, Instagram |
| People in Research | Website |
| Postural Tachycardia Syndrome UK (PoTS UK) | Twitter, Facebook |
| PsAZZ Support Group | Contacted network |
| Scleroderma & Raynaud’s UK (SRUK) | Twitter, Facebook |
| The Erythromelgalgia Warriors | Website, Twitter, Facebook |
| UK Gout Society | Twitter |
| Vocal | Contacted network |
