## Additional File 3 for "What do people living with chronic pain want from a pain forecast? A research prioritisation study"

Additional File 3: Planned structure of the focus group

| Section of focus group | Purpose | Example questions/statements | Anticipated duration (mins) |
| --- | --- | --- | --- |
| Introduction and general overview | - Greeting - Use of Zoom - Review of ground rules - Ice breaker | - Zoom: how to use hands up function - We sent around some ground rules as suggested by the university. Is there anything that is missing that you would like to add? | 20 |
| Introduction to our ideas | - Outline the context of research - Explain proposed research | Presentation | 5 |
| Breakout rooms | - Understand initial thoughts about the research and perceived interest in different predictands | - Here are some common patterns of pain severity. Which one(s) do you relate to? - What pain features would you want to know about? | 15 |
| Break |  |  | 10 |
| Group discussion | - Bring thoughts from breakout rooms together | - What pain features did you come up with? - Were there any in other groups that you think are good that you hadn’t thought of? | 15 |
| Building a questionnaire | - Outline of a questionnaire that would be meaningful | - We want to build a questionnaire to ask other people what information they would like to know about pain patterns in the future. Here are some example questions that we have thought of - Are there any questions here that you think we should remove? - Are there any questions that we should include? - What possible answers should we have? - What information would you add or remove to the images to make them clearer? | 20 |
| Conclusion | - Thank participants & let them know what the next steps are |  | 5 |
