## Additional File 4 for "What do people living with chronic pain want from a pain forecast? A research prioritisation study"

CENTRE FOR EPIDEMIOLOGY VERSUS ARTHRITIS

MANCHESTER 1824 The University of Manchester

CLOUDY WITH A CHANCE OF PAIN

#### Info and consent

Thank you for your interest in helping us build a pain forecast. In 2016, the Centre for Epidemiology Versus Arthritis at the University of Manchester ran a large study, collecting data about people's symptoms and comparing this to the weather. You can find out more about this study by visiting the Cloudy with a Chance of Pain website.

Using this data, we are now aiming to build a pain forecast. This will be like a weather forecast but used for predicting pain severity in the near future. Being able to forecast pain severity in this way will provide benefits for patients in the future.

During this survey we are interested in learning about the information that you would be interested in receiving about your pain severity in the near future.

The next page will outline the information that we will collect in this study, your data protection rights and how to find out more information. If you are still happy to continue, you will be asked a series of questions about our research.

Thank you for your time.

Before you continue, it is important that you read the following information to understand what this research entails and what will happen to your information by completing this questionnaire. If you have any further questions, please use the contact details provided at the end of this page.

The research is being conducted by Claire Little (PhD student, Centre for Epidemiology Versus Arthritis, University of Manchester) and supervised by Professor John McBeth (Centre for Epidemiology Versus Arthritis, University of Manchester). It has been approved by the University of Manchester Proportionate Ethic Committee, [UREC reference number 2021-11862-19751].

To participate in this research, you must be over the age of 18, with a chronic pain condition, live in the UK, and be able to read English.

It is up to you whether you want to take part. On the next page, you consent to us using your anonymous answers for research purposes. During the questionnaire, you may stop at any time and your responses will not be used. However, once you submit the questionnaire, it will not be possible to remove your responses from the anonymised aggregated data.

Your responses will only be viewed by researchers at the University of Manchester. We will use these to understand your views on a pain forecast in our future work. It is also possible that some answers may be published or used in a student thesis. By continuing, you consent to this use.

Please note the following information in relation to the processing of your data.

- The University of Manchester, as Data Controller for this project takes responsibility for the protection of the personal information that this study is collecting about you. In order to comply with the legal obligations to protect your personal data the University has safeguards in place such as policies and procedures. All researchers are appropriately trained.

- Data will be held securely by the research team on behalf of the University of Manchester according to the University's data protection and information security policies.
- Your responses to this questionnaire will be stored for 5 years, in accordance with the University's Record Retention Policy. We are collecting and storing this personal information in accordance with the General Data Protection Regulation (GDPR) and Data Protection Act 2018 which legislate to protect your personal information. The legal basis upon which we are using your personal information is "public interest task" and "for research purposes".
- For more information about the way we process your personal information and comply with data protection law please see our Privacy Notice for Research Participants.

Complaints Procedure

If you have any complaints, you can contact Claire's supervisor, Professor John McBeth at

If you wish to make a formal complaint to someone independent of the research team or if you are not satisfied with the response you have gained from the researchers in the first instance then please contact The Research Governance and Integrity Officer, Research Office, Christie Building, The University of Manchester,

Oxford Road, Manchester, M13 9PL,
or by telephoning 0161 306 8089.

Contact details
If you have any further questions, please contact Claire Little at.

Have you read the information on the previous page? (Required)

- Yes
- No

Do you confirm that you are over 18, have a chronic pain condition, live in the UK and can read English? (Required)

- Yes
- No

Do you understand that participation in this study is voluntary but once you have submitted the questionnaire, it will not be possible to remove your answers from the data set? (Required)

- Yes
- No

Do you provide consent for aggregated responses to be used in a student thesis, reports, presentations or journals? (Required)

- Yes
- No

#### Demographic

What is your gender? (Required)

- Male
- Female
- Non-binary / third gender
- Prefer not to say

What is your age? (Required)

- 18-25
- 26-35
- 36-45
- 46-55

- 56-65
- 65+
- Prefer not to say

Has your doctor ever told you that you have any of the following conditions? (Required; Choose as many as relevant)

- Rheumatoid Arthritis
- Osteoarthritis
- Spondyloarthropathy
- Gout
- Unspecific Arthritis
- Fibromyalgia
- Chronic headache
- Neuropathic pain
- Other (please specify)

Where do you generally experience pain? (Required; Choose as many as relevant)

- Mouth or jaw
- Neck or shoulder
- Back
- Stomach or abdominal
- Hip

- ☐ Knee
- ☐ Hands
- ☐ Feet
- ☐ Pain at multiple sites
- ☐ Pain all over body

#### Block 2

We know that people's pain severity can vary every day. In 2016, we collected information about people's pain severity on a daily basis, ranging from no pain to very severe pain.

Here is an example. For the first few days, the person has reported no pain, but on day 5, they begin reporting very severe pain. Later on, the pain severity reduces to moderate and then mild.

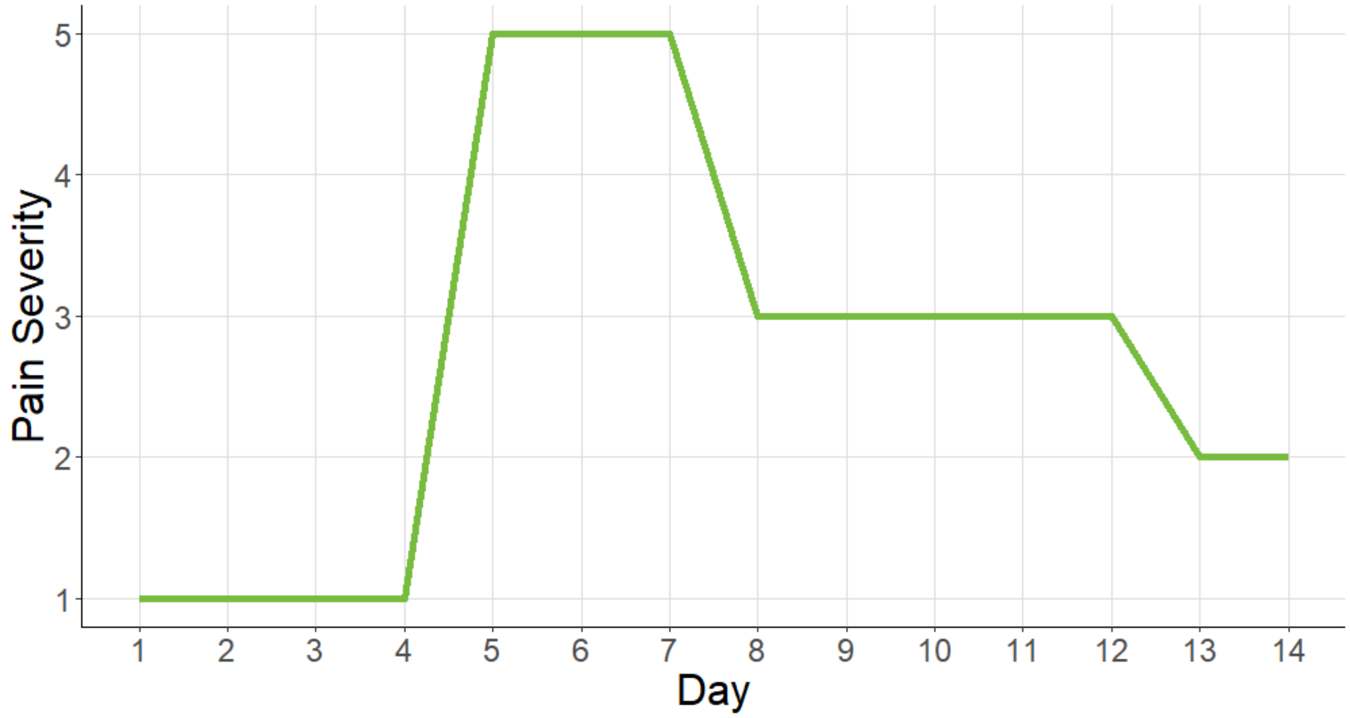

We are hoping to use this information to produce a pain forecast, that could predict pain severity in the future. We are interested in your opinions about this pain forecast.

**Block 10**

#### Section 1: How do you experience pain?

Everyone experiences pain in different ways. In this section, we are interested in understanding your experience of different pain patterns.

Some people experience pain patterns as shown by the graphs below. Other people do not experience any of these pain patterns. Which, if any, of these pain patterns do you ever experience? (Choose, by clicking on the graphs, as many as required)

☐ Constant pain that doesn't tend to change

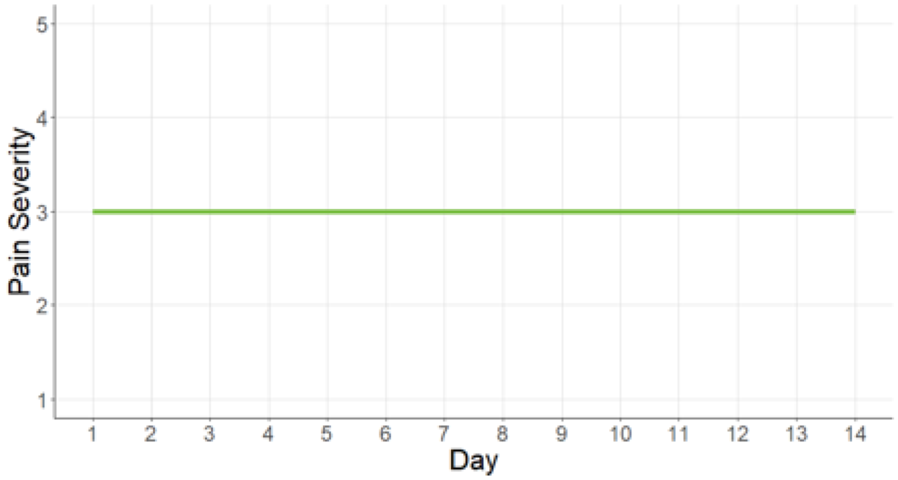


☐ Rapid increase in pain severity that lasts a few days (pain flare)

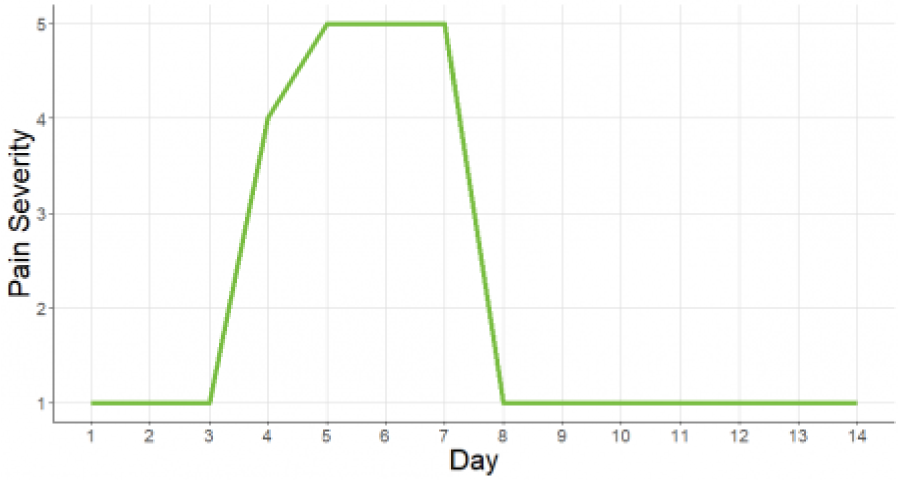


☐ Generally high pain with a period of lower pain

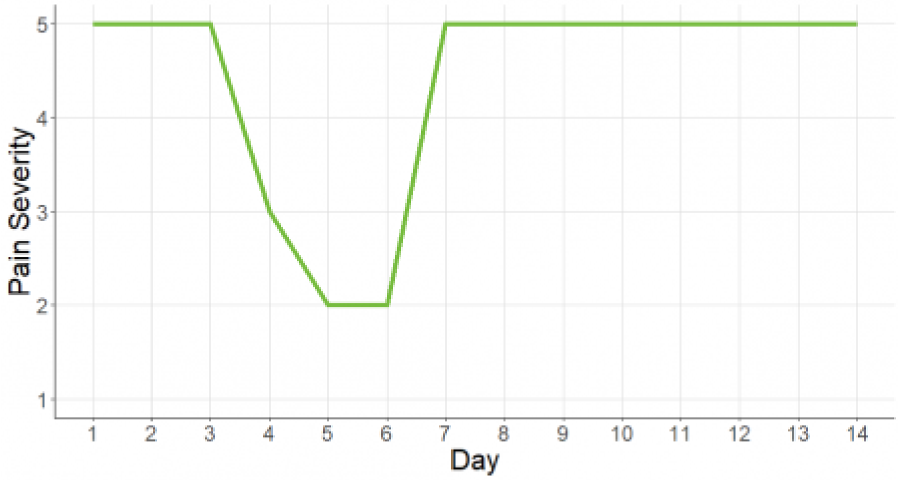

☐ Variable pain severity that changes day-to-day

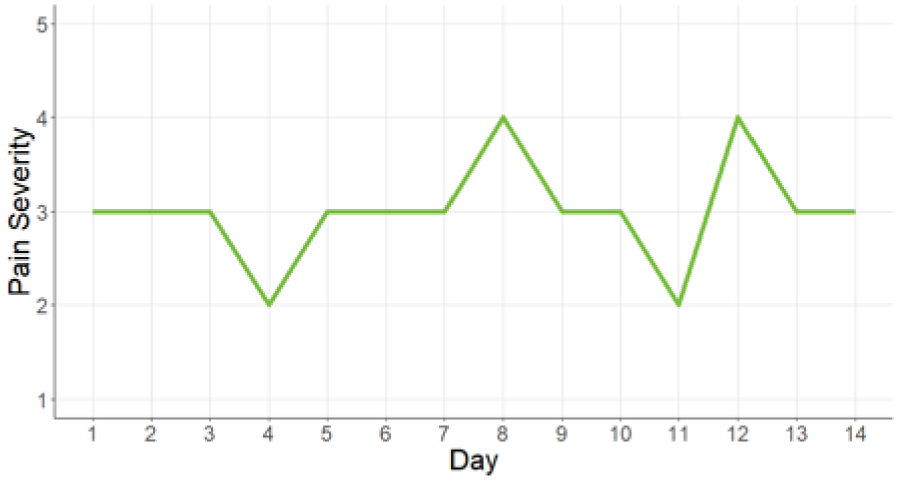


☐ Occasional increases in pain severity that last a very short time (e.g. one day)

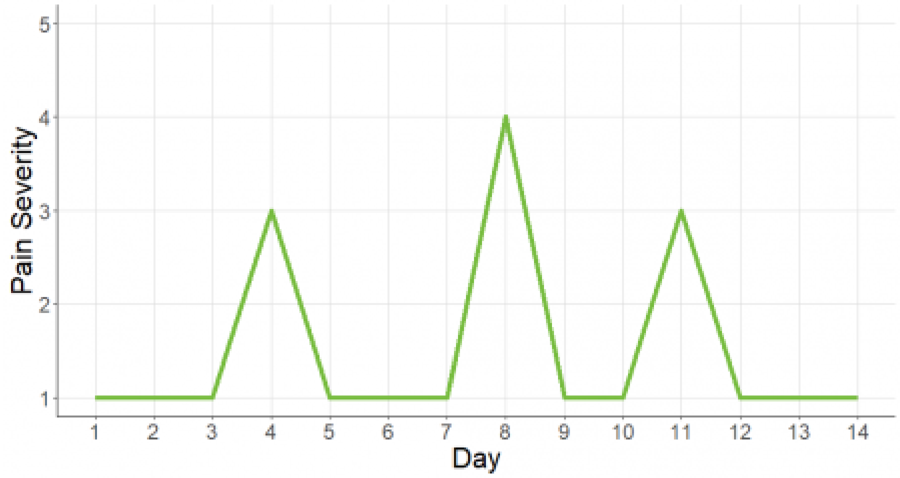


☐ Occasional decreases in pain severity that last a very short time (e.g. one day)

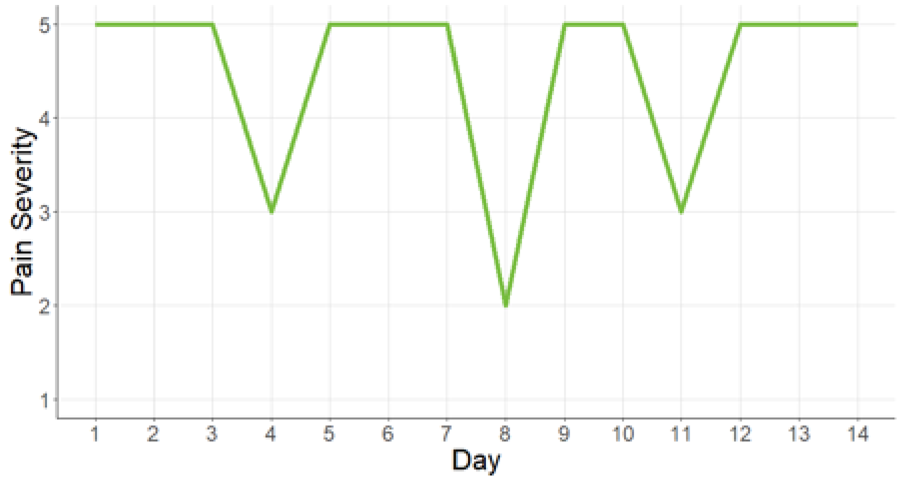


☐ I experience a different pain pattern. (Please describe below).

☐ None of these represent pain that I experience

### Section 2: What information do you want from a pain forecast?

We are using pain patterns (like the ones on the previous page) to build a **pain forecast**, that would predict your pain severity in the future. We are interested in understanding the information that you would like from a pain forecast.

To start with, please think very generally about the type of information that you would like from a pain forecast. Which of the following would you like a pain forecast to provide for you? (Choose as many relevant)

☐ Information about the fluctuations in pain severity (e.g. how stable pain severity will be over the week)

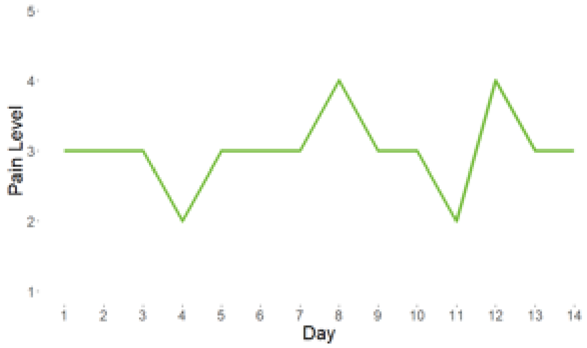


☐ Information about pain severity on a scale of 1 (no pain) to 5 (very severe pain) (e.g. what pain severity will be on this scale tomorrow)

☐ Information about a period of low/no pain severity (e.g. timing/duration)

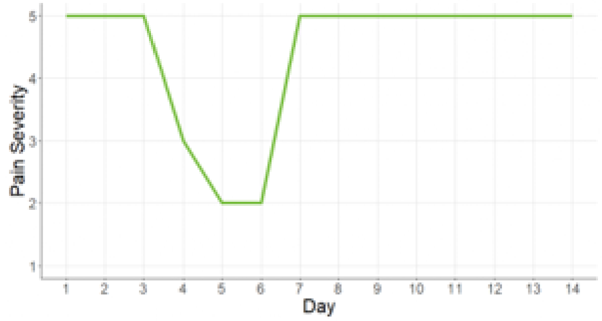


☐ Information about a pain flare (e.g. timing/duration/severity)

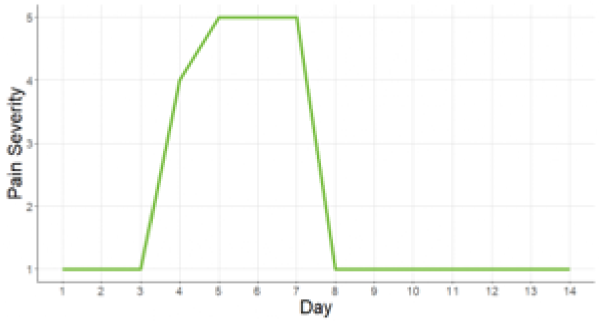


☐ [ ] Other:

If we could predict a period of low pain severity, what specific information would you want to know? (Please rank the responses from most wanted information to least wanted information by dragging the responses to be in the correct order)

When might the period of low pain severity start?

When might the period of low pain severity end?

How many days might the period of low pain severity last for?

Might my pain-related quality of life also be better?

Might my other symptoms also be better? (e.g. fatigue, morning stiffness, sleep quality)

Is there any other information about periods of low pain severity that you would like to know about? (Optional)

If we could predict a pain flare, what specific information would you want to know? (Please rank the responses from most wanted information to least wanted information, by dragging the responses to be in the correct order)

When might the pain flare start?

When might the pain flare end?

How many days might the pain flare last for?

Might my pain-related quality of life be lower?

Might my other symptoms vary? (e.g. fatigue, morning stiffness, sleep quality)

How severe will the pain flare be?

Is there any other information about pain flares that you would like to know about? (Optional)

**Block 4**

#### Section 3: How would you use a pain forecast?

Now that you have considered the type of information that a pain forecast could provide, we are interested in how you would use a pain forecast.

On the previous page, we asked you to consider information that you might want from a pain forecast. If a pain forecast could provide useful information for you, do you think that you would use a pain forecast?

- Definitely yes
- Probably yes
- Might or might not
- Probably not
- Definitely not

If so, what would you use a pain forecast for? (Choose as many as required)

- ☐ To know when my pain severity might be better/worse
- ☐ To understand the triggers of my pain
- ☐ To help choose which medication to take
- ☐ To help plan non-pharmacological interventions (i.e. other methods of helping pain that aren't medication)
- ☐ To help make plans (e.g. shopping, social )
- ☐ Other

Powered by Qualtrics
