## Additional File 5 for "What do people living with chronic pain want from a pain forecast? A research prioritisation study"

Additional File 5 – Sensitivity analysis

In this appendix, we acknowledge that a large proportion of our respondents reported fibromyalgia (46%) and osteoarthritis (33%) as a chronic pain condition. We conducted sensitivity analyses to compare (1) the responses between those respondents who reported fibromyalgia as a pain condition and those that did not, and (2) the responses between those respondents who reported osteoarthritis as a pain condition and those that did not.

| **Table S1: Responses to the question: "Which of the following would you like a pain forecast to provide for you?" Participants could select more than one option.** | | | |
| --- | --- | --- | --- |
| **Pre-specified response** | **Number (and percentage) of full population who selected response** | **Number (and percentage) of subgroup with fibromyalgia who selected response** | **Number (and percentage) of subgroup with osteoarthritis who selected response** |
| Information about a pain flare | 100 (67.6%) | 47 (69.1%) | 34 (69.4%) |
| Information about fluctuations in pain severity | 94 (63.5%) | 47 (69.1%) | 27 (55.1%) |
| Information about pain severity on a scale of 1 to 5 | 70 (47.3%) | 30 (44.1%) | 23 (46.9%) |
| Information about a period of low/no pain severity | 51 (34.5%) | 25 (36.8%) | 15 (30.6%) |
| Other/None | 13 (8.8%) | 5 (7.4%) | 5 (10.2%) |

For each question, we report the number and percentage of respondents in the disease subgroups who selected each response and perform a chi-squared test to test whether these responses are significantly different from these not in the corresponding subgroup.

Results to the question: "Which of the following would you like a pain forecast to provide for you?” are reported in Table S1. A chi squared test of the responses for this question from participants who did not report fibromyalgia against those who did report fibromyalgia gave a p-value of 0.2414. For the same question, a chi squared test of the responses from participants who did not report osteoarthritis against those who did report osteoarthritis gave a p-value of 0.2202. Therefore, there is no evidence that the subgroups gave statistically significantly different responses to the population.

| **Table S2: Responses to the question: “If a pain forecast could provide useful information for you, do you think that you would use a pain forecast?”** | | | |
| --- | --- | --- | --- |
| **Response** | **Number (and percentage) of full population who selected response** | **Number (and percentage) of subgroup with fibromyalgia who selected response** | **Number (and percentage) of subgroup with osteoarthritis who selected response** |
| Definitely not | 0 (0%) | 0 (0%) | 0 (0%) |
| Probably not | 11 (7.4%) | 2 (2.9%) | 4 (8.2%) |
| Might or might not | 24 (16.2%) | 9 (13.2%) | 8 (16.3%) |
| Probably yes | 63 (42.6%) | 27 (39.7%) | 16 (32.7%) |
| Definitely yes | 50 (33.8%) | 30 (44.1%) | 21 (42.9%) |

Results to the question: "If a pain forecast could provide useful information for you, do you think that you would use a pain forecast?" are reported in Table S2. A chi squared test of the responses from participants who did not report fibromyalgia against those who did report fibromyalgia gave a p-value of 0.2133. For the same question, a chi squared test of the responses from participants who did not report osteoarthritis against those who did report osteoarthritis gave a p-value of 0.2133. Therefore, there is no evidence that the subgroups gave statistically significantly different responses to the population.

| **Table S3: Responses to the question "What would you use a pain forecast for?" Participants could select more than one option.** | | | |
| --- | --- | --- | --- |
| **Pre-specified response** | **Number (and percentage) of full population who selected response** | **Number (and percentage) of subgroup with fibromyalgia who selected response** | **Number (and percentage) of subgroup with osteoarthritis who selected response** |
| To help make plans (e.g. shopping, social) | 123 (83.1%) | 59 (86.8%) | 45 (91.8%) |
| To understand the triggers of my pain | 113 (76.4%) | 53 (77.9%) | 38 (77.6%) |
| To know when my pain severity might be better/worse | 92 (62.2%) | 46 (67.6%) | 28 (57.1%) |
| To help plan non-pharmacological interventions | 70 (47.3%) | 29 (42.6%) | 22 (44.9%) |
| To help choose which medication to take | 46 (31.1%) | 24 (35.3%) | 21 (42.9%) |
| Other/None | 16 (10.8%) | 6 (8.8%) | 5 (10.2%) |

The responses to the question: "What would you use a pain forecast for?" are reported in Table S3. A chi squared test of the responses from participants who did not report fibromyalgia against those who did report fibromyalgia gave a p-value of 0.2243. For the same question, a chi squared test of the responses from participants who did not report osteoarthritis against those who did report osteoarthritis gave a p-value of 0.2243. Therefore, there is no evidence that the subgroups gave statistically significantly different responses to the population.
